# Understanding barriers to effective injury care in low-income contexts: a mixed methods analysis of insights from medical trainees and traffic law enforcement first responders in Uganda

**DOI:** 10.1101/2025.02.22.25322712

**Authors:** Herman Lule, Benson Oguttu, Micheal Mugerwa, Michael Lowery Wilson, Jussi P. Posti

**Affiliations:** Department of Surgery, Kiryandongo Regional Referral Hospital, Kigumba, Uganda; Injury Epidemiology and Prevention (IEP) Research Group, Turku Brain Injury Centre, Department of Clinical Neurosciences, Turku University Hospital and University of Turku, Turku, Finland; Center for Health Equity in Surgery and Anesthesia, Institute of Global Health Sciences, University of California, San Francisco, USA; Department of Public Health, Botswana-Havard Partnership, Gaborone, Botswana; Department of Biostatistics and Public Health, California Baptist University, California, USA; Heidelberg Institute of Global Health (HIGH), University Hospital and University of Heidelberg, Heidelberg, Germany; Neurocenter, Department of Neurosurgery and Turku Brain Injury Center, Turku University Hospital and University of Turku, Turku, Finland

**Keywords:** Injury Care, Barriers to Access, Low-Income Countries, Trauma Management, Healthcare Systems

## Abstract

**Background:** Injury-related mortality exhibits a significant social gradient, particularly in low-middle-income countries (LMICs), with approximately 4.5 million premature deaths annually.

**Objective:** This study explores prehospital and in-hospital barriers to timely injury care as perceived by frontline healthcare personnel in Uganda.

**Methods:** We utilized a mixed methods approach, gathering qualitative data from five hundred frontline workers including surgical residents, interns, and law enforcement professionals, alongside quantitative data from 1,003 trauma patients. Barriers were categorized into pre- and in-hospital trauma team-related, patient-related, and systemic healthcare challenges.

**Results:** From frontline workers, prehospital barriers included delays in emergency medical services activation (21.2%), ambulance arrival (19.3%), and transportation to hospitals (15.2%) whereas in-hospital barriers primarily involved supply shortages (28.3%), delays in identifying life-threatening injuries (27.3%), and insufficient critical care services (26.3%). Among the 1003 audited trauma patients, 41.5% (n=416) faced barriers during treatment. The most common barriers were delays in treatment decisions (n=232, 23.1%) and securing necessary supplies (n=180, 17.9%). Presence of a barrier correlated with higher odds of unfavourable Glasgow Outcome Scale scores compared to those without barriers [OR 1.750, 95% CI (1.497-2.047) vs. OR 0.556, 95% CI (0.436-0.708), p<0.001]. Moreover, the odds of mortality were higher for those whom a barrier was encountered compared to those where there was no barrier [OR 1.901, 95% CI (1.057-3.420) vs. OR 0.588, 95% CI (0.397-0.869), p<0.001].

**Conclusions:** Our findings highlight the need for multifaceted targeted interventions, integrating frontline healthcare perspectives to improve trauma care delivery in LMICs which face both prehospital and in-hospital disparities to accessing injury care.

## INTRODUCTION

Injury related mortality is characterised by significant social gradients [1]. Globally, the burden of injury is disproportionately high in nations with poor economies, contributing to approximately 4.5 million premature deaths annually [2], [3], [4].

Barriers to trauma prevention and care drive inequalities in injury-related deaths and disability-adjusted life years (DALYs) across both low- and high-income countries. For instance, research highlights substantial regional disparities in trauma-related mortality rates, particularly in Europe, where Eastern Europe experiences rates nearly three times higher than those in Western Europe [1], [2]. In the United States, compelling evidence demonstrates desperate trauma outcomes that are linked to disparities in ethnicity and insurance coverage where uninsured persons of low socioeconomic status are the most vulnerable to adverse injury outcomes [5] whereas in Australia, rural geographical disparities to injury care are well documented [6].

Conversely, whereas recent trends show declining inequalities in injuries amongst high income countries [2], in low- and middle-income countries (LMICs), socioeconomic inequities, inadequate infrastructure, and shortages of human resources have continued to exacerbate these challenges, resulting in disparities in access to injury care both between and within regions, for instance in cities versus rural regions [7], [3].

In Africa, emergency medical services have poor coverage in the majority of LMICs [8]; [9]. For instance, despite high trauma volumes, notable barriers have been identified in countries such as Rwanda and Uganda, including inadequate insurance coverage and limited trauma care resources ranging from lack of ground ambulances to a limited number of trauma care specialists and rigid non-decentralized referral systems [10], [11].

However, existing studies often lack a comprehensive analysis of the barriers faced at the individual patient level, which are influenced by geographical and cultural contexts [10], [12], [11]. In Uganda, studies which have attempted to investigate some of these barriers have limited their scope to pre-hospital factors mainly in urban settings [13]; [14], and have excluded insights from frontline health care workers. This phenomenon can be attributed, in part, to the challenges associated with acquiring information from key frontline stakeholders. These challenges arise from their demanding work schedules, remote geographical locations, privacy concerns, and the aspiration to manage the heterogeneity of research participants, which often proves to be realistically unattainable.

To effectively address these complex issues, mixed methods research is essential for understanding the specific barriers to timely injury care in low-income country settings, particularly from the perspectives of healthcare personnel involved in initial patient contact. Prior research has identified mixed methods as particularly suitable for investigating complex healthcare systems [15]. In their recent systematic review, Whitaker et al. have recommended that future research assessing barriers to injury care should utilize innovative combined methodologies to investigate both prehospital and in-hospital delays, as well as the associated care processes and outcomes [12].

Consequently, the aim of this study was to identify the prehospital and in-hospital barriers to timely injury care as perceived by personnel who are the first point of contact and are actively engaged in the care of injury patients in a low-income country context, as well as to elucidate how these barriers impact injury outcomes. We structured the identified barriers into two frameworks, focusing on both prehospital and in-hospital challenges, while examining both infrastructural and human resource factors. Additionally, we aimed to delineate the specific barriers encountered at both the individual-patient and trauma-team level during the real-time execution of treatment.

## MATERIALS AND METHODS

### Study design

This exploratory sequential mixed methods study was nested within a broader clinical trial which investigated the impact of medical trainee and traffic law enforcement led trauma teams and training to patient outcomes (Pan African Clinical Trial Registry: PACTR202308851460352) [16]. We fitted the two-framework approaches into the concept of three temporal delays in accessing surgical care within the context of LMICs including seeking, reaching and receiving as previously described by Vaca et al. [17]. This theoretical framework permitted the integration of qualitative and quantitative data to gain a deeper understanding of obstacles to injury care. The research is documented in accordance with good reporting of mixed methods studies guidelines [18].

### Study settings

The study was conducted from September 2019 to August 2023 at six regional tertiary hospitals in Uganda: Jinja, Hoima, Fort Portal, Mubende, Kiryandongo, and Kampala International University Teaching Hospital which also function as internship training centers for surgery, trauma, and emergency medicine. The trauma and casualty units are staffed by multidisciplinary teams of consultant surgeons, residents, interns, and clinical medical students. Trauma patients typically arrive via ground ambulances, traffic police patrols or public transport, where they are assessed and treated by specialist trainees, interns, or medical students, who may refer cases to consultants based on complexity.

### Study population and source of data

Our exploratory sequential approach began with qualitative followed by quantitative data collection. First, qualitative data were collected from surgical residents, interns (doctors and nurses), and third- and fifth-year medical students associated with the surgery departments of the participating hospitals. Additionally, we engaged law enforcement professionals specializing in road traffic regulations, who serve as first responders to road traffic-related injuries within the municipalities that constitute the catchment areas of these hospitals. The qualitative data were collected through self-administered semi-structured questionnaires featuring open-ended options. These questionnaires were administered as part of a pre-test conducted prior to the development training of rural trauma teams previously described for the aforementioned frontline health workers [19], [20].

In contrast, quantitative data were gathered from investigator-administered survey questionnaires to patients who sustained motorcycle-related injuries, received care at any of the participating hospitals during the study period and participated in the motorcycle trauma outcome registry (MOTOR trial project) [16], [21].

### Sample size estimation and sampling methods

The qualitative component involved 500 trauma care frontliners (434 medical and 66 traffic law enforcement professionals). The qualitative sample was estimated using a hypergeometric formula, based on the annual number of trainee medical professionals at the respective teaching hospitals and the average number of traffic law enforcement officers deployed at the police headquarters serving those hospitals, as previously reported [19], [20]. In contrast, the quantitative component assessed barriers to injury care among 1,003 trauma patients treated at the six study sites, whose sample size was derived from an open-source R Shiny application for the MOTOR registry trial, which was a cluster randomized, parallel controlled trial with discrete time decay correlation structures for multiple periods as detailed in Lule et al. [16].

### Eligibility criteria

The qualitative component comprised trauma care frontliners including surgery chief residents, heads of medical interns, traffic police officers stationed on major highways leading to the hospitals and clinical-year medical students participating in surgical and traumatology rotations during the study period. The quantitative component involved motorcycle crash patients aged 2 to 80 years who presented to the emergency departments of six study sites during the same period. More detailed inclusion and exclusion criteria are documented in the study protocols [16], [19].

### Statement of ethics approval and consent to participate

Ethical approval was secured from the Research and Ethics Committees of Mbarara University of Science and Technology (Ref: MUREC 1/7; 05/5-19) and the Uganda National Council for Science and Technology (Ref. No. SS 5082). All study participants provided written informed consent prior to their involvement in the study.

### Data collection

Data were collected to identify barriers presumed to contribute to delays in accessing injury care, as indicated by existing literature [12]. The data collection process was conducted in two sequential phases. In the initial phase, surgery interns, clinical year students, and law enforcement professionals completed a semi-structured survey questionnaire to assess and quantify barriers to prehospital delays. This phase was conducted during a one-day consultative meeting as part of patient-public engagements at each data collection site in September 2019, prior to the MOTOR trial project [16]. Participants ranked prehospital delays using a six-point Likert scale, where one indicated “not a barrier” and six indicated “very strong barrier.” Additionally, medical interns provided their insights on in-hospital team and system barriers, employing a four-point Likert scale ranging from 1 (“least important barrier”) to 4 (“very important barrier”). An open-ended qualitative component was included to assess law enforcement perspectives on the integration of emergency evacuation teams on highways and the existence of “rural networks of first lay responders” in the regions under study, aiming to elucidate potential barriers to prehospital care and inform the establishment of a pilot rural trauma team and registry focused on motorcycle-related injuries.

In the subsequent quantitative phase, a prospective motorcycle trauma outcome data registry (MOTOR trial project) was established from September 2019 to August 2023 [16]. Consultants, surgical residents, and interns involved in trauma care documented barriers encountered during providing definitive injury treatment for 1,003 registered patients at the study sites. Data were collected using paper-based survey questionnaires, which were subsequently coded and uploaded to the secure REDCap electronic system [22], managed by the University of Turku. Each documented barrier triggered an alert, prompting an audit of the patient’s case file during weekly meetings. These open discussions aimed to elucidate the nature of the barriers, with verbatim and anonymous documentation of responses from both those who reported the barriers and those directly involved in patient care. This comprehensive approach aimed to capture a nuanced understanding of the barriers affecting injury care access in real-time to enhance the effectiveness of future interventions.

### Data analysis

The data from this mixed-methods study were analyzed independently, remaining concealed from the MOTOR trial team until the analysis of the primary clinical trial findings was completed. It was anticipated that the insights gained from this mixed-methods investigation would enrich and inform the interpretation of the trial outcomes.

In the initial phase (qualitative analysis), concerning prehospital barriers (delay 1), participants could select from six potential responses, with scores ranging from a minimum of 1 to a maximum of 6. A barrier category was deemed significant if it achieved an overall mean score exceeding the hypothesized average of 3.5. For in-hospital team barriers (delay 2) and system barriers (delay 3), there were four potential responses, with scores ranging from 1 to 4. A barrier category was considered important if it surpassed the hypothesized average of 2.5. The overall ranking of barriers was determined by calculating weighted mean scores, derived from the sum of individual Likert component scores multiplied by the number of responses for each component, subsequently divided by the valid sample size. This methodology has been previously validated for assessing barriers to trauma care in LMICs [12]. For example, in relation to prehospital delays, responses were scored on a Likert scale as follows: 1 (not a barrier), 2 (very weak barrier), 3 (weak barrier), 4 (averagely weak barrier), 5 (strong barrier), and 6 (very strong barrier).

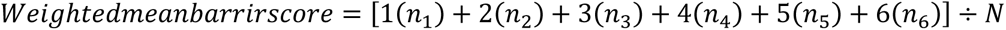

Where *n*_*i*_= number of people responding per component, N=sample size

In the second phase (quantitative analysis), responses were categorized into four distinct groups: (a) no barriers encountered in the execution of definitive injury care; (b) team-related barriers that lead to delays in the activation, identification, prioritization, recognition, and timely referral of skilled emergency teams for life-threatening injuries; (c) individual barriers, whether from patients or clinicians, that contributed to delays in decisions regarding surgery, initiation of treatment, and referral of injuries exceeding local capacity; and (d) in-hospital system barriers that cause delays in securing essential supplies, including oxygen, anesthetics, sutures, blood products, and access to functional laboratory and imaging diagnostics, as well as theatre space and intensive and critical care services. These categories were subsequently represented as proportions. The mean rank scores for each individual barrier across various cadres (third, fifth, intern, police) were compared to the hypothesized mean using the non-parametric Kruskal-Wallis equality of proportions rank test. Normal distribution was assessed using Levene’s test.

### Data integration

Mixed-effects regression analyses were performed in SPSS 20.0 to determine the correlation between barrier and injury outcomes including mortality and morbidity at 90-days post-injury, as determined by the Glasgow outcome scale [23], and trauma expectation factor (TEFS)/trauma outcome measure (TOMS) scores [24]. A description of how these tools were used to evaluate morbidity associated with neurological and musculoskeletal injuries including assessments for pain, physical function, disability, satisfaction with treatment, and overall quality of life is detailed in the study protocol [16]. Directed content analysis of themes derived from the transcribed qualitative data were conducted in (NVivo 14, release 2023) and visualized at wordcloud.com.

## RESULTS

Of the 500 trauma care frontliners who participated in semi-structured interviews for qualitative analysis, there were 135 (27.0%) females and 365 (73.0%) males of which were 66 (13.2%) traffic law enforcement officers and 434 (86.8%) were medical trainees. Of the 1003 trauma patients in the motorcycle trauma outcome (MOTOR) registry for quantitative analysis, there were 817 (82%) males and 186 (19%) females. The overall median age (IQR) was 28 (22-37) years. The sociodemographic characteristics of both care providers and patient participants have been detailed elsewhere [20][preprint].

### Quantitative findings

#### Phase 1: Analyses of barriers to injury care based on medical trainee and traffic law enforcement frontliners

##### Prehospital barriers (Delay 1)

The three highest-ranked barriers to accessing injury care identified in the assessment of prehospital factors were as follows: delays in the activation and mobilization of emergency medical services (21.2%), delays attributable to prehospital response times, including the arrival of ambulances at the scene (19.3%), and prolonged transportation times to the hospital (15.2%) (see Fig. 1).

**Figure 1:**
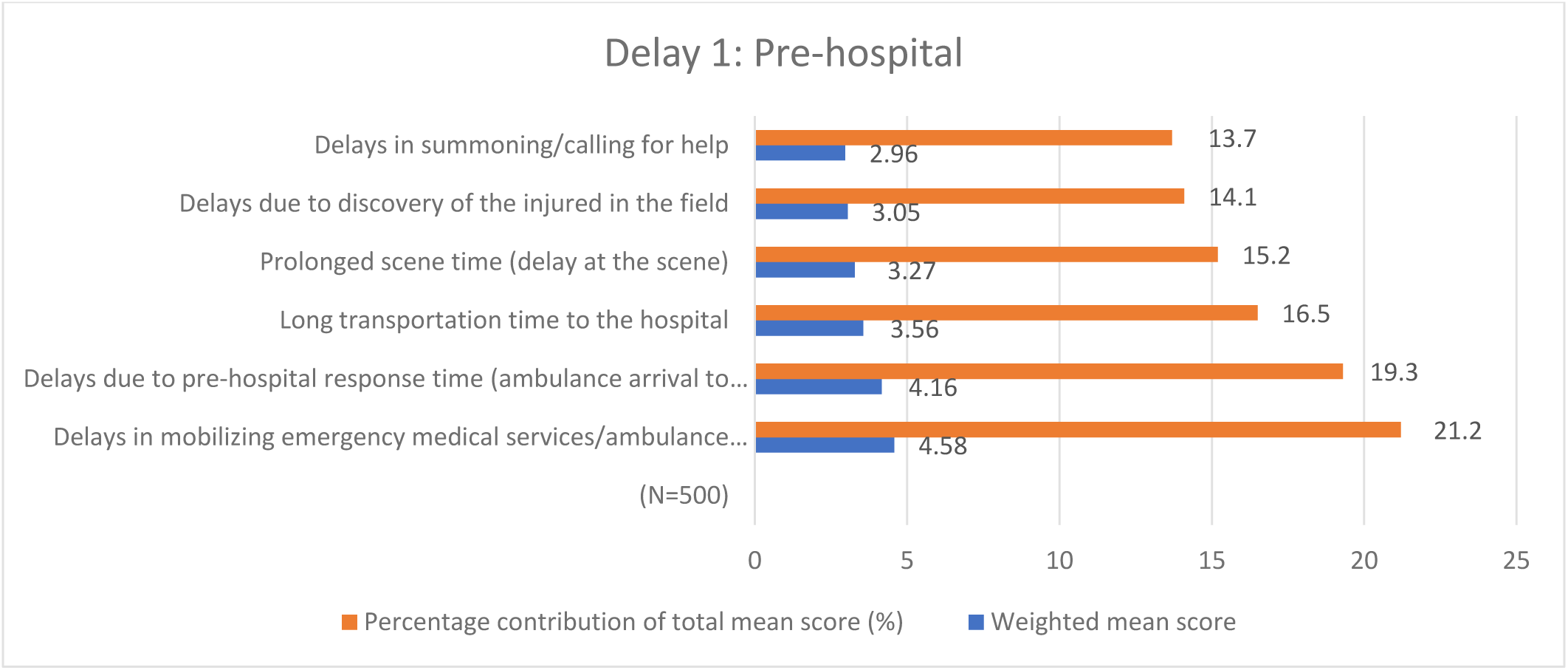
showing overall weighted mean scores for prehospital barriers (Delay 1) ranked from bottom-up

##### Team Barriers (Delay 2)

Delay in identification of life threatening injuries (27.3%) and referral to higher trauma centers (26.1%) were the most cited in-hospital team barriers (Fig.2).

**Figure 2:**
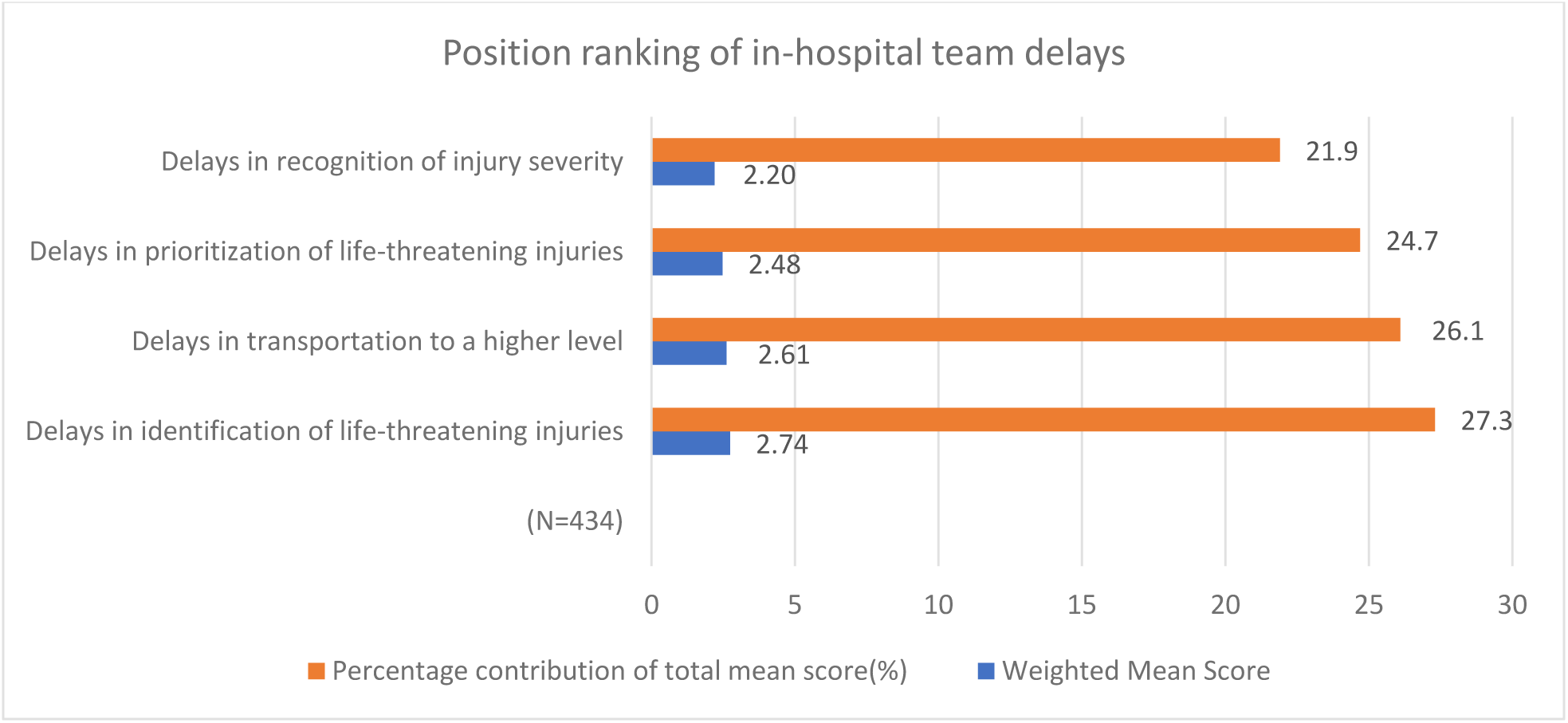
overall mean weighted scores for in-hospital team barriers (Delay 2)

##### In-hospital system barriers (Delay 3)

The most cited in-hospital system barriers to injure care were lack of supplies (28.3%) and critical care services (26.3%) (Fig. 3).

**Figure 3:**
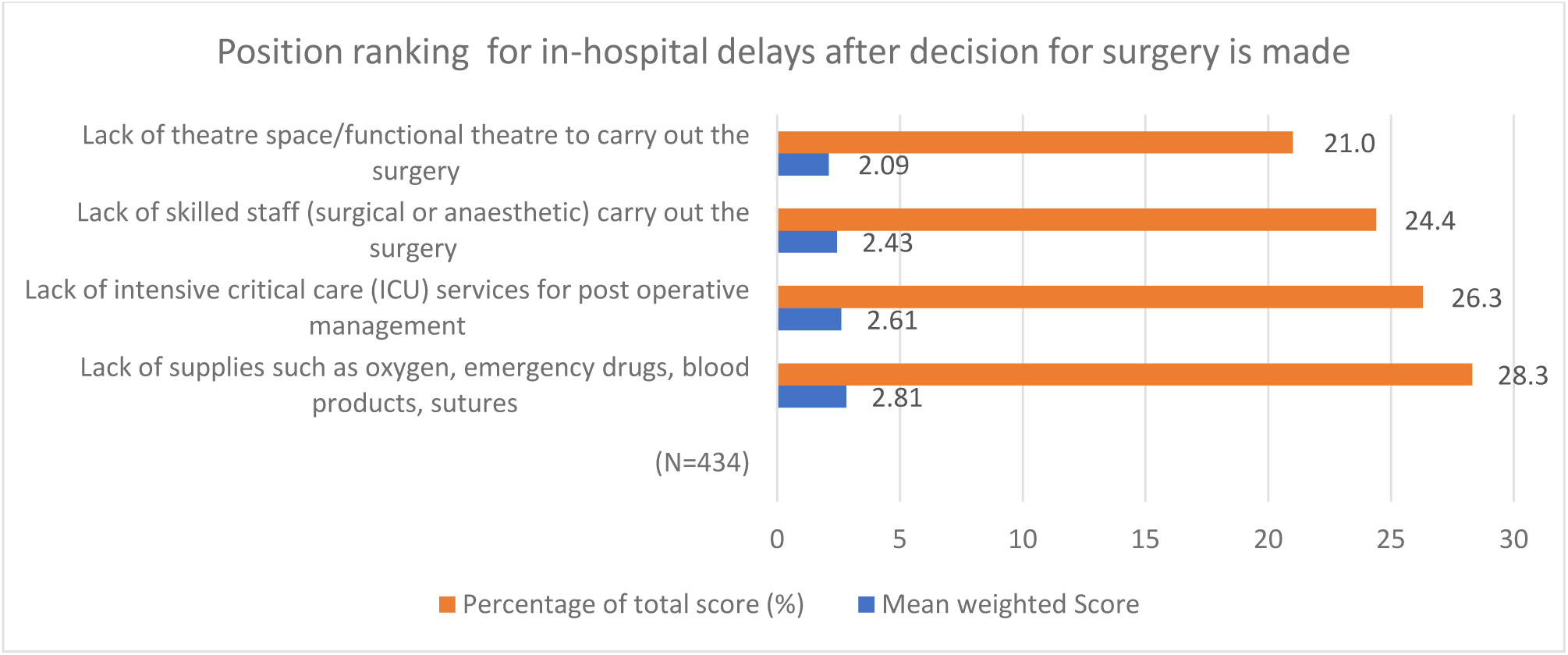
Mean weighted scores for in-hospital barriers after decision to operate (Delay 3)

The Kruskal-Wallis test showed statistically significant differences in how the various cadres ranked the various delays (Table 1).

**Table 1:**
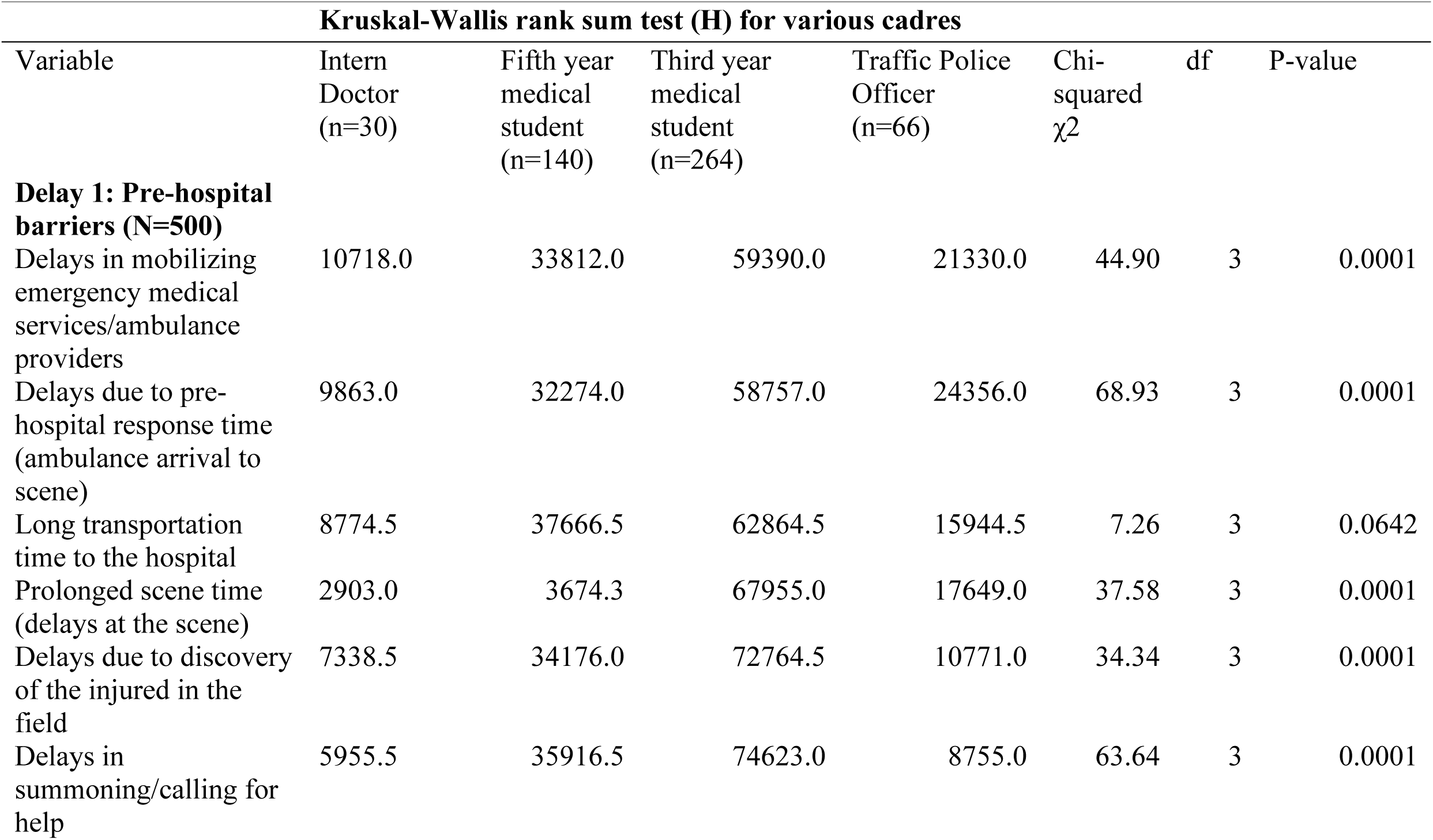
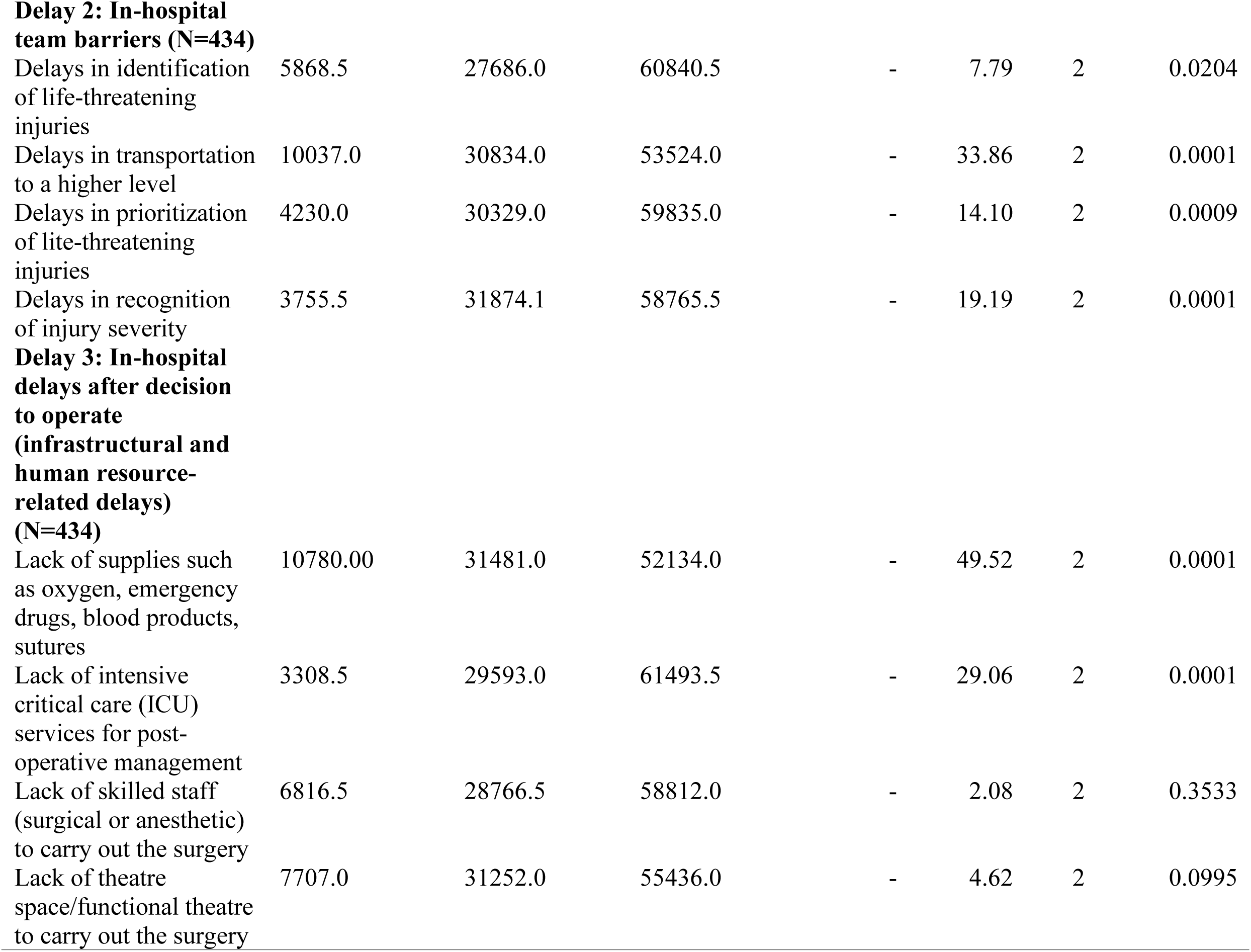
Shows the mean rank scores for different barriers by the various cadres.

| Kruskal-Wallis rank sum test (H) for various cadres |  |  |  |  |  |  |  |
| --- | --- | --- | --- | --- | --- | --- | --- |
| Variable | Intern Doctor (n=30) | Fifth year medical student (n=140) | Third year medical student (n=264) | Traffic Police Officer (n=66) | Chi-squared $\chi^2$ | df | P-value |
| **Delay 1: Pre-hospital barriers (N=500)** |  |  |  |  |  |  |  |
| Delays in mobilizing emergency medical services/ambulance providers | 10718.0 | 33812.0 | 59390.0 | 21330.0 | 44.90 | 3 | 0.0001 |
| Delays due to pre-hospital response time (ambulance arrival to scene) | 9863.0 | 32274.0 | 58757.0 | 24356.0 | 68.93 | 3 | 0.0001 |
| Long transportation time to the hospital | 8774.5 | 37666.5 | 62864.5 | 15944.5 | 7.26 | 3 | 0.0642 |
| Prolonged scene time (delays at the scene) | 2903.0 | 3674.3 | 67955.0 | 17649.0 | 37.58 | 3 | 0.0001 |
| Delays due to discovery of the injured in the field | 7338.5 | 34176.0 | 72764.5 | 10771.0 | 34.34 | 3 | 0.0001 |
| Delays in summoning/calling for help | 5955.5 | 35916.5 | 74623.0 | 8755.0 | 63.64 | 3 | 0.0001 |
| **Delay 2: In-hospital team barriers (N=434)** |  |  |  |  |  |  |  |
| Delays in identification of life-threatening injuries | 5868.5 | 27686.0 | 60840.5 | - | 7.79 | 2 | 0.0204 |
| Delays in transportation to a higher level | 10037.0 | 30834.0 | 53524.0 | - | 33.86 | 2 | 0.0001 |
| Delays in prioritization of life-threatening injuries | 4230.0 | 30329.0 | 59835.0 | - | 14.10 | 2 | 0.0009 |
| Delays in recognition of injury severity | 3755.5 | 31874.1 | 58765.5 | - | 19.19 | 2 | 0.0001 |
| **Delay 3: In-hospital delays after decision to operate (infrastructural and human resource-related delays) (N=434)** |  |  |  |  |  |  |  |
| Lack of supplies such as oxygen, emergency drugs, blood products, sutures | 10780.00 | 31481.0 | 52134.0 | - | 49.52 | 2 | 0.0001 |
| Lack of intensive critical care (ICU) services for post-operative management | 3308.5 | 29593.0 | 61493.5 | - | 29.06 | 2 | 0.0001 |
| Lack of skilled staff (surgical or anesthetic) to carry out the surgery | 6816.5 | 28766.5 | 58812.0 | - | 2.08 | 2 | 0.3533 |
| Lack of theatre space/functional theatre to carry out the surgery | 7707.0 | 31252.0 | 55436.0 | - | 4.62 | 2 | 0.0995 |

#### Phase 2: In-hospital audit of barriers to injury care for patient participants based on their various attending clinicians

In the study of 1,003 audited trauma cases, our findings indicated that only 58.5% (587/1,003) received treatment for their injuries without their clinical teams encountering any barriers in the provision of definitive care. In contrast, the remaining patients faced one or more hindrances at the individual-patient, trauma-team, or hospital-system level (see Fig. 4).

**Figure 4:**
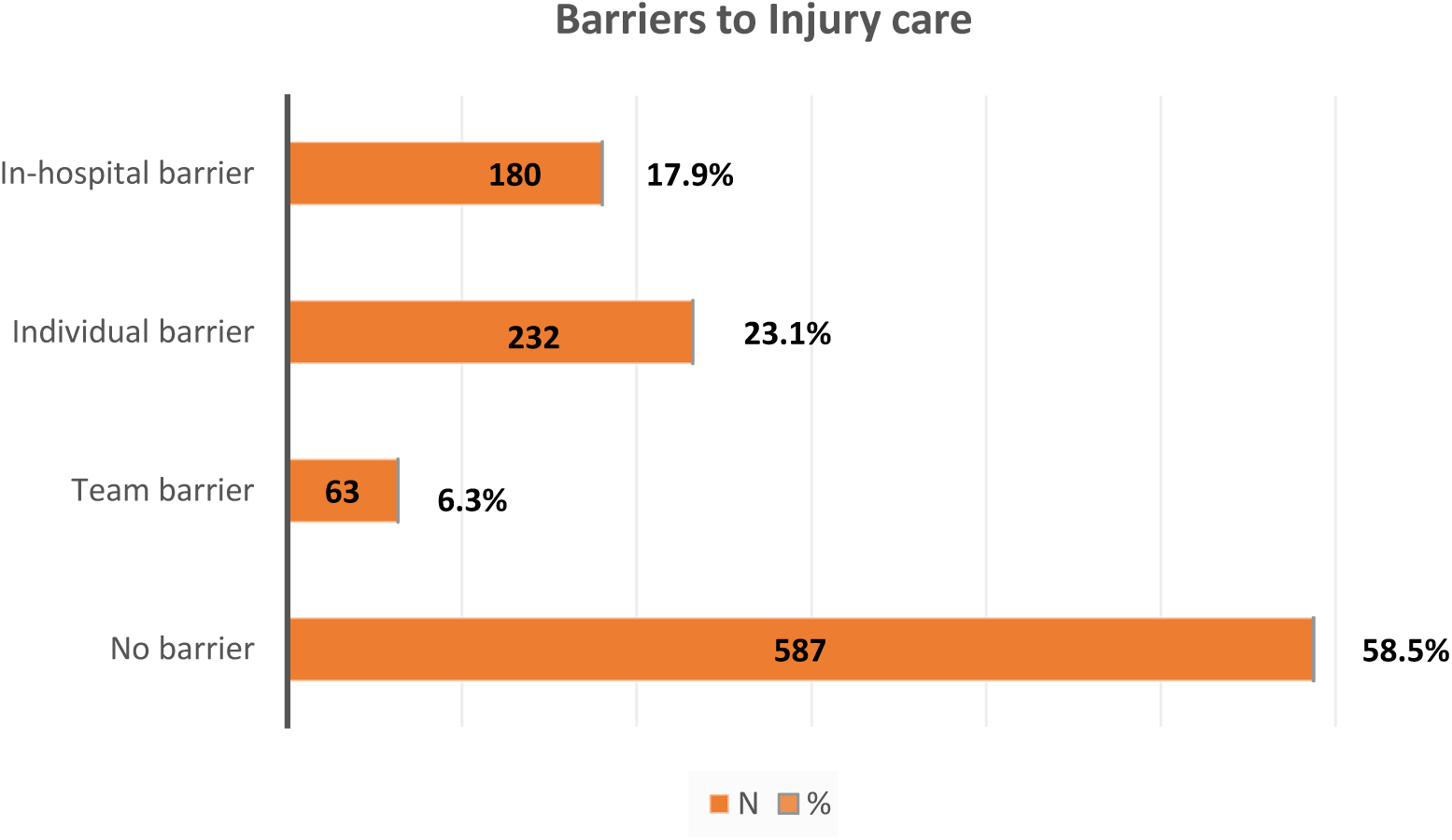
Barriers to definitive injury care based on an audit of 1003 trauma cases at six regional referral hospitals.

Additionally, complete 90-day follow-up data were analyzed for 887 of the 1,003 individuals with traumatic brain injuries. After adjusting for injury severity based on the Glasgow Coma Scale (GCS), the odds of an unfavorable outcome on the Glasgow Coma Outcome Scale were significantly higher for those who encountered barriers (Odds Ratio [OR] 1.750, 95% Confidence Interval [CI] 1.497-2.047) compared to those who did not encounter barriers (OR 0.556, 95% CI 0.436-0.708), p < 0.001. Furthermore, the odds of mortality were also elevated for patients who faced barriers compared to those without barriers (OR 1.901, 95% CI 1.057-3.420 vs. OR 0.588, 95% CI 0.397-0.869), p < 0.001. Among the 805 individuals with complete follow-up for musculoskeletal injuries, 195 exhibited unfavorable trauma outcomes (as measured by TOMS at the 90-day follow-up being less than TEFS at baseline). However, the presence of barriers did not significantly influence patient-reported outcomes for musculoskeletal injuries (TEFS-TOMS) (OR 0.963, 95% CI 0.691-1.341, p = 0.821).

### Qualitative findings

First, we found that there were no designated rural networks of emergency rescue police to intervene for traffic crash victims.

> *“The role of evacuation of traffic crash victims is not specifically designated to particular officers within our jurisdiction which means any road traffic officer on duty can handle such cases” said one of the law enforcement officials.*

In addition, limited human resource capacity was reported as a key obstacle for the law enforcement professionals to participate in pre-hospital evacuation of injured patients.

> *“Often we are understaffed and not able to undertake transfer responsibilities timely as this would mean leaving one’s station vacant which would not be acceptable in our line of duty”, said one of the law enforcement officials.*

Furthermore, a lack of key focal persons concerned with emergencies at recipient hospitals and suboptimal communication technologies were cited as key barriers to timely injury care.

> *“Of course, you want to communicate timely but the poor communication technologies and network breakdown will let you down. Our intercom is not directly linked to that of the hospital emergency departments unless one is chanced to have a physical contact there. Nevertheless, we are required to drop them (patients) by the government emergency hospital departments”*

Lastly, there were also concerns of bias and mistrust which led to non-disclosure of injury-related information which could delay the emergency evacuation process.

> *“We would be delighted to have an integration of emergency evacuation rescue services that is well coordinated along with the health ministry, I guess this would increase our public trust. You want to help evacuate these accident victims, but they think you are interrogating, arresting, and prosecuting them”.*

### Convergence of phase one and phase two

The analysis revealed a convergence of key themes regarding perceived barriers to injury care, as reported by frontline health workers who completed semi-structured questionnaires in the initial consultative phase, and those reported by attending clinicians during the real-time execution of patient treatment in the subsequent phase. The identified barriers predominantly included delays in emergency evacuation and late hospital arrival from the trauma scene (prehospital), primarily attributed to a shortage of ground ambulances and the reliance on public transport. Additionally, there was a notable lack of resources for both pre- and post-operative critical care, as well as limited diagnostic and infrastructural capabilities, such as functional CT scans, which contributed to delays in treatment decisions. Furthermore, a general shortage of essential supplies, including pharmaceuticals and blood products, was observed. A summary of the key themes derived from the qualitative analyses of open-ended questions concerning barriers to injury care is presented in Figure 5.

> *“We wanted to obtain head and brain CT scans but when the service in our government national referral hospital was down, the patient’s next of kin delayed mobilizing financial resources to outsource one from private. We don’t have direct referral pathways to private hospitals so then we had to wait until patients’ relatives availed us with the results of the investigations before operating” Said an attending neurosurgeon.*

> *“He arrived late in a taxi as it appeared there was no fuel in the ambulance from the referring hospital. The staff there say were understaffed and were unable to accompany the patient as no one would cover their duty. Our ICU is full though, so we plan to refer him to a private hospital, but we are facing difficulties with direct communication and his family is still mobilizing resources for ICU costs. He doesn’t seem to be insured”. Said an attending anesthesiologist.*

> *“As an intern doctor, my role was to stabilize the patient before the theatre once the decision to operate was made by the consultant. It frustrated me when I could not get the lab results and blood products on time because of having no reagents yet my boss was on my neck”*

**Figure 5:**
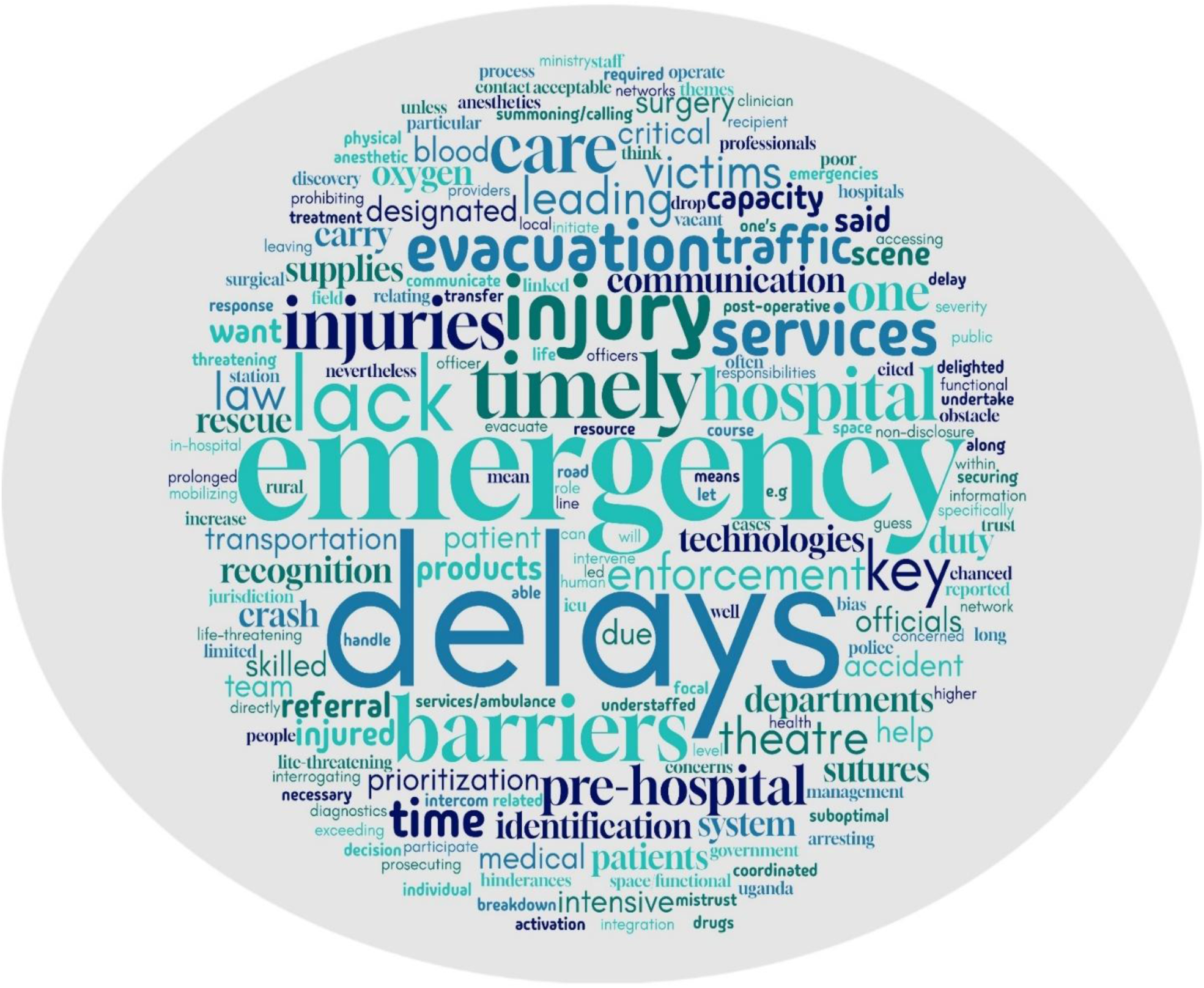
Word cloud summarizing key themes emerging from qualitative analyses of open ended questions

## DISCUSSION

This study aimed to identify prehospital and in-hospital barriers to injury care as reported by personnel who are the first point of contact and are actively involved in the management of injured patients. The findings from the study reveal critical barriers in both the prehospital and in-hospital management of trauma patients, highlighting systemic inefficiencies that compromise patient outcomes. The prehospital barriers, primarily delays in emergency medical services activation (21.2%) and ambulance arrival (19.3%), reflect significant gaps in the emergency response framework. These delays can have dire consequences, as they prolong the time to definitive care and potentially worsen the severity of injuries. Moreover, the lack of designated rural networks for emergency response, as indicated by law enforcement officials, exacerbates the situation in less urbanized areas, where timely access to emergency services is crucial. The absence of clear protocols for traffic crash victim evacuation further complicates the response, leading to a reliance on any officer available rather than trained personnel, which may negatively impact the quality of care provided. Similar barriers have been earlier identified in rural settings of other LMICs [12], [25].

Qualitative analyses shed light on the human factors contributing to these barriers, particularly the limitations faced by law enforcement professionals and referring clinicians. The reported understaffing and lack of designated roles for emergency road traffic response personnel indicate a need for strategic workforce management and clear delineation of responsibilities. This issue is compounded by inadequate communication technologies, which hinder effective coordination between emergency responders and hospitals. The inability to establish timely communication not only delays patient transfer but can also result in critical information being lost, further jeopardizing patient care [10]. Therefore, addressing these infrastructural shortcomings is essential for improving the overall effectiveness of trauma care systems, particularly in remote settings.

Furthermore, the study highlights the psychological barriers to effective emergency response, such as bias and mistrust among the public towards law enforcement officials. This mistrust can result in non-disclosure of injury-related information, which is crucial for timely intervention and settlement of insurance claims linked to injuries to mitigate delays due to out of pocket payments. The concerns voiced by law enforcement officials regarding public perceptions of their role during emergencies indicate a need for increased community engagement and educational initiatives aimed at building trust. Integrating police services with healthcare systems could enhance coordination and alleviate public fears, ultimately leading to better outcomes for trauma patients [26]. As such, a comprehensive approach that encompasses both operational and human elements is necessary to address the multifaceted barriers identified in this study.

In-hospital barriers present equally alarming challenges, with supply shortages (28.3%) and delays in identifying life-threatening injuries (27.3%) being particularly prevalent. These findings underscore the need for robust logistical frameworks within hospitals to ensure that critical supplies are consistently available, and that staff can quickly assess, diagnose, and treat life-threatening conditions. The correlation between the presence of barriers and unfavourable Glasgow Outcome Scale scores (OR 1.750) further emphasizes that these systemic issues not only hinder immediate care but also have lasting effects on patient recovery and survival. Additionally, the increased odds of mortality (OR 1.901) among those facing barriers highlight the urgent need for interventions aimed at streamlining care processes and resource allocation within trauma care systems. Studies conducted in Rwanda, Ghana, and South Africa have assessed the application of multiple data collection methods to identify context-specific barriers to injury care that contribute to mortality and disability, and our findings are consistent with these studies [25].

Lastly, the barriers identified by attending clinicians during the real-time execution of treatment, such as inadequate diagnostic resources, transportation failures, financial challenges, and resource shortages, highlight systemic issues within the healthcare framework. In a recent Delphi study conducted in Malawi [27], the absence of readily available physical resources emerged as the most significant factor contributing to in-hospital delays. Conversely, the high financial cost of care and the unaffordability of emergency transport were identified as the primary factors leading to prehospital delays [27]. These challenges reflect inadequate resource allocation to trauma care, not only in LMICs but also in HICs. A systematic review of barriers to injury care in 21 countries across North and South America found that resources and equipment constituted 58% (33/57) of barriers, followed by the absence of protocols at 51%, understaffing at 30%, transport and logistics challenges at 28%, and financial limitations at 26% [28]. Research from Australia and Europe elaborates on these challenges, highlighting rural-urban and socio-economic disparities amid declining social support structures [29], [30]. To mitigate these barriers, investment in reliable diagnostic equipment and partnerships with private facilities for CT scans are essential. Improving ambulance logistics to ensure sufficient staffing and fuel supply, along with establishing clear referral pathways and financial support systems for uninsured patients, can enhance patient transport and continuity of care. Additionally, optimizing the procurement of medical supplies and developing robust supply chain management will ensure timely access to necessary resources for healthcare providers.

### Policy implications

Our study has identified significant barriers to injury care. The results indicate multiple barriers present at various stages of the trauma care continuum. Most of these identified barriers are applicable to other low-middle-income countries [12], [25], [27]. Consequently, the barriers to injury care from the perspective of first responders must be addressed to accurately conceptualize their expected roles. To ensure favorable outcomes, it is essential to tackle the three delays through the collaboration of multidisciplinary teams, focusing on safe and timely transportation, quality pre-hospital resuscitation, and comprehensive post-crash care. Stakeholders, including the Ugandan Ministry of Health, Regional Police Commanders and National Road Safety Council, should engage in planning meetings to address these critical issues.

In the context of individual-patient and trauma-team delays, the interplay between inadequate insurance coverage and low disposable incomes often results in significant delays as patients’ relatives seek to mobilize funds for necessary investigations. Consequently, attending clinicians are frequently left to make surgical decisions based solely on clinical signs and symptoms. For example, one notable barrier arises from patients needing to secure resources for essential diagnostic procedures, such as a brain CT scan, which can cost approximately $130, an amount that is substantial relative to the average monthly salary of green-collar workers in Uganda [17]. The pursuit of diagnostic precision through gold standard investigations is essential for ensuring high-quality surgical outcomes. Furthermore, the apprehension regarding potential medical-legal litigation further complicates clinicians’ decision-making processes, particularly as patients increasingly assert their autonomy and seek greater involvement from their families in treatment decisions.

In-hospital system delays in injury care were primarily attributed to a lack of supplies and critical care services, with these barriers ranking second in an audit of patient handling. Key challenges included securing emergency supplies such as oxygen, anesthetics, and blood products, as well as limited access to operating theatres and intensive care units. Previous studies in Uganda have highlighted a shortage of essential equipment for trauma resuscitation. A recent investigation by Ningwa et al. [10] found inadequate availability of crucial medical devices, including blood pressure monitors, defibrillators, and electrocardiograms, along with a lack of permanent staff in 73% of casualty units surveyed. Additionally, a Delphi study by Whitaker et al. [12], identified inadequate trauma care coordination, financial barriers, and a lack of essential resources as significant obstacles to receiving timely injury treatment in LMICs. The dual deficiencies in equipment and human resources imply severe hinderance to life-saving interventions, contributing to a substantial proportion of preventable trauma deaths in LMICs.

### Study strengths and limitations

One notable strength of the study is its mixed methods approach, which enhances the depth of analysis and allows for a nuanced exploration of barriers to injury care. Additionally, by centering on the perspectives of those directly involved in patient care, the study provides valuable insights into real-world challenges encountered in LMIC settings. However, there are limitations that should be acknowledged. First, the use of Likert scales with fewer than four items may pose challenges for interpretation, and while the study employs parametric tests to enhance statistical power, this approach might not align with best practices for exploratory research that lacks prior hypotheses [31]. Furthermore, the study assumes that accumulated Likert items can serve as a reasonable surrogate for continuous interval measurements. Although this assumption facilitates the computation of overall mean scores and objective rankings for barriers, it may potentially compromise the precision of central tendency measures [32]. Lastly, while we endeavored to interpret the data objectively through triangulation and use of parametric tests, we acknowledge the possibility of bias arising from the integration of diverse data paradigms that employed varying collection and sampling methods. This concern reflects a significant and ongoing debate within the field of mixed methods research [33].

## Conclusion

This mixed methods study has evaluated the key barriers to injury care within the context of frontline health workers in low resource settings. We identified significant barriers in both prehospital and in-hospital trauma care that adversely affect patient outcomes. Systemic inefficiencies, including delays in emergency response and inadequate resources, underscore the urgent need for improved logistical frameworks and clear referral protocols. Additionally, addressing the human factors, such as staffing challenges, public mistrust and out of pocket payments for investigations, is crucial for enhancing coordination between emergency services and healthcare facilities to enable timely treatment decisions. A comprehensive strategy that integrates operational improvements with community engagement is essential to mitigate these barriers and optimize trauma care delivery. Future studies should be robust multicenter prospective cohorts to investigate how these barriers impact the long-term treatment outcomes of injured patients in diverse socioeconomic settings.

## Supporting information

Ethical Approval and GRAMMS Checklist

## Acknowledgements

The authors wish to express their gratitude to the staff at various medical referral institutions in Uganda, including Jinja, Mbarara, Hoima, Mubende, Kiryandongo, and Kampala International University Teaching Hospitals. Their provision of facilities was instrumental in facilitating the data collection for this study. Furthermore, the authors acknowledge the study participants for their willingness to dedicate their time to participating in this investigation.

## Author contributions

HL was the lead researcher who conceived, designed, and implemented the study, and wrote the initial draft of the manuscript. HL and BO assisted with administrative tasks, data acquisition and participant follow-up. HL and MM were responsible for data analysis and cross-validation. MLW and JPP supervised the study, lobbied for funding and provided critical review of the final manuscript. All authors read and approved the final version of the manuscript.

## Ethical approval

Ethical approval for this research involving human participants was obtained from the Research and Ethics Committees of Mbarara University of Science and Technology (Reference: 1/7; 05/5-9) and the Uganda National Council for Science and Technology (Reference: SS 5082).

## Data availability statement

The data collection tools and anonymized datasets pertaining to this study and the parent trial (Pan African Clinical Trial Registry: PACTR202308851460352), are publicly accessible [16], [20], [21].

## List of abbreviations

GOS: Glasgow Outcome Scale
HICS: High-Income Countries
LMICs: Low-Middle-Income Countries
TEFS: Trauma Expectation Factor Score
TOMS: Trauma Outcome Measure Score

