## Supplementary material for "Understanding barriers to effective injury care in low-income contexts: a mixed methods analysis of insights from medical trainees and traffic law enforcement first responders in Uganda": Ethical Approval and GRAMMS Checklist: Good reporting of a mixed methods study GRAMMS Checklist.docx

| **Good reporting of a mixed methods study (GRAMMS) checklist^⸸^** | **Section: (Page)** |
| --- | --- |
| Describe the justification for using a mixed methods approach to the research question | Introduction: pg 3  Study design: pg 4  Data collection: pg 5,6  Data analysis: pg 6 |
| Describe the design in terms of the purpose, priority and sequence of methods | Study design: pg 4  Data collection: pg 5,6  Data analysis: pg 6 |
| Describe each method in terms of sampling, data collection and analysis | Data source: pg 4,5  Sample size estimation: pg 5  Data collection: pg 5,6  Data analysis: pg 7 |
| Describe where integration has occurred, how it has occurred and who has participated in it | Data analysis: pg 6  Data integration: pg 7 |
| Describe any limitation of one method associated with the present of the other method | Study strengths and limitations: pg 14 |
| Describe any insights gained from mixing or integrating methods | Study strengths and limitations: pg 14 |
| ^⸸^O'Cathain A, Murphy E, Nicholl J. The quality of mixed methods studies in health services research. J Health Serv Res Policy. 2008;13: 92-98. | |
